# Functional assessment improves discrimination of clinical and biomarker-defined Alzheimer’s disease

**DOI:** 10.64898/2026.07.29.26359236

**Authors:** Kalliopi Mavromati, Austin Jon Dibble, Lucie Tvrda, Connor Dalby, Jack D. Beazer, Lynne Hughes, Sean P. Kennelly, Terence J. Quinn

**Affiliations:** School of Cardiovascular and Metabolic Health, University of Glasgow, Glasgow, UK; School of Psychology and Neuroscience, University of Glasgow, Glasgow, UK; Institute of Memory and Cognition, Tallaght University Hospital, Dublin, Ireland; Department of Medical Gerontology, School of Medicine, Trinity College Dublin, Dublin, Ireland; Global Alzheimer’s Platform Foundation, Washington, USA

**Keywords:** Alzheimer’s disease, functional assessment, risk algorithms, blood biomarkers, older adults

## Abstract

**INTRODUCTION:** Accurately identifying risk of Alzheimer’s Dementia (AD) is essential for supporting people living with symptoms in clinical settings, as well as recruiting adults in prospective medical research. Various algorithms have been created to calculate AD risk based on evidenced risk factors which are weighted toward a total score. As daily life conditions determining risk change at scale, it remains unclear how effective gold standard algorithms remain in modern cohorts.

**METHODS:** In the Bio-Hermes-001 diverse cohort, we assessed algorithm discrimination and calibration in six outcomes: classifying AB PET binary outcome (negative N = 603, positive N = 342); phosphorylated tau-217 binary outcome (pTau-217 negative N = 166, positive N = 469); participants with Healthy Cognition (N = 417) from probable AD (N = 272); HC from Mild Cognitive Impairment (N = 312), HC from pooled MCI or AD; and MCI from AD. Approximately a third of the cohort are individuals from populations typically underrepresented in dementia research (HC: 19%; MCI: 24%; AD: 33%).

**RESULTS:** Hosmer-Lemeshow tests and Brier score demonstrate acceptable calibration of all algorithms except the oldest algorithm. However, Receiver Operating Characteristic (ROC) curves and the associated area under the curve (AUC) estimates evidenced that in this cohort only the BDSI exceeded conventional thresholds for good discrimination (.8 AUC in HC-AD classification, with AUC approximately .7 in the other clinical, AB PET, and pTau-217 comparisons). When the functional item is removed from the BDSI score, it remains acceptably calibrated, but DeLong tests reflect statistically significant reduction in discriminatory performance for all group comparisons. The two earliest published algorithms were only chance-level accurate.

**DISCUSSION:** In a contemporary, diverse cohort, most established dementia risk algorithms had limited power in discriminating amyloid positivity, pTau-217 positivity, and current cognitive status despite acceptable calibration. Including a functional measure markedly improved discrimination across both clinical and biomarker-defined outcomes, suggesting that proximal indicators of cognitive vulnerability are critical for identifying individuals with underlying AD-related pathology.

## Introduction

Quantifying risk for neurodegenerative disease, including Alzheimer’s Disease (AD), is central to population-level planning, clinical screening, and the design of trials for early intervention or prevention. Over the past two decades, multiple algorithms have been developed to identify individuals at elevated risk for AD and dementia based on combinations of demographic, clinical, and lifestyle factors[1].

Inherently grounded in the populations and historical contexts in which they are developed, risk algorithms consist of risk determinants reflecting shared conditions and experiences, including educational opportunity, occupation and lifestyle patterns[1]. The impact and relevance of these risk factors fluctuate over time[2] as political and economic developments shape lived experience at the community and individual level[1] and as historically underrepresented populations (URP) increasingly comprise the age groups disproportionally affected by dementia[3].

An early epidemiological risk model that became the gold standard, Cardiovascular Risk Factors, Aging, and the Incidence of Dementia (CAIDE[4]) centred cardiometabolic health in quantifying dementia risk. Subsequent tools including Lifestyle for BRAin Health (LIBRA[5, 6]), Australian National University AD Risk Index (ANU-ADRI[7, 8]) and the Cognitive Health and Dementia Risk Index (CogD) with its AD-specific version (COGD-AD[8]), expanded this framework to incorporate lifestyle and psychosocial indicators as well as co-occurring conditions. These developments reflected growing recognition of the cumulative nature of brain health risk, which is shaped by health behaviours and individuals’ circumstances across the life course.

However, behaviour does not occur in a vacuum. Behaviour change requires capability, opportunity, and motivation[9]; hence the feasibility of truly modifying lifestyle risk factors is shaped by broader socioeconomic and environmental context[10–12]. Consequently, markers such as BMI that are distal to lived experience may not uniformly reflect modifiable lifestyle risk across populations or as environments in which individuals live continue to evolve over time.

Moreover, comprehensive operationalisation of these algorithms for use in existing real-world datasets is often constrained by data availability and granularity for individual risk factors[13]. For example, datasets may not contain detailed information on weekly cognitive activity or fish intake required for a target risk score. This necessitates evidence that risk algorithms retain clinical utility even where data are incomplete or operationalised using imperfectly aligned proxies[7, 14, 15], such as alcohol consumption recorded only as rare or frequent behaviour where a risk score requires weekly unit intake. Given the heterogeneity of approaches used to capture health behaviours across individual studies, partial implementation may disproportionately affect scores that rely on distal lifestyle or environmental exposures, which are often less consistently operationalised across datasets[13, 16, 17].

By contrast, proximal indicators of cognition and function commonly used in health research are typically assessed using standardised measures and may therefore offer more robust insight into individual vulnerability. A notable exception is the Brief Dementia Screening Indicator (BDSI[18]) that includes a single self-reported item assessing difficulty with managing money or medications as part of a short list of items to support clinical decision to refer individuals for dementia screening. Whether this functional component meaningfully contributes to contemporaneously discriminatory performance, particularly relative to other established algorithms, remains unclear.

To that end, we aimed to explore how existing risk algorithms would distinguish clinical classification subgroups in the uniquely diverse Bio-Hermes-001 cross-sectional cohort[19]. For this exploratory secondary analysis, we reviewed existing scores that could be implemented in the dataset and compared their capacity to discriminate cognitive status and AD pathology burden measured via amyloid beta PET scan positivity. We further explored discrimination of positivity in blood-based pTau-217, a clinically scalable biomarker available in the cohort that is increasingly used to identify AD-related pathology[20]. In a post hoc ablation analysis, we investigated which risk factors best accounted for the discrimination and calibration of the best performing score.

## Methods

### Cohort

We accessed the Bio-Hermes-001 dataset[19] through the Data Challenge[21]. The sample of N = 1001 was representative of community dwelling adults aged between 60 and 85 years. Recruitment dedicated resource to involving individuals from URP in dementia research. Screening and blood sampling was undertaken during the first visit, with follow-up Amyloid Beta (AB) PET scan where possible. Investigators of the original study classified participants as cognitively normal (CN), mild cognitive impairment (MCI) or mild Alzheimer’s disease (AD) following National Institute on Aging – Alzheimer’s Association[22] criteria (see Supplemental Materials for thresholds in Mini Mental State Exam[23], Ray Auditory Verbal Learning Test[24] and Functional Activities Questionnaire[25], Table S1). Ethical approval for use of this dataset in this was provided under the Data Challenge.

### Risk scores

Following a review of relevant literature, we identified 7 risk scores that could be calculated in the Bio-Hermes-001 dataset: Cardiovascular Risk Factors, Aging, and the Incidence of Dementia (CAIDE[4]) as previously adapted to include APOE-e4 allele carriership[26]; Lifestyle for BRAin Health (LIBRA[5, 6]) using the Geriatric Depression Scale for depression[27] [28]; Brief Dementia Screening Indicator (BDSI[18]) with the deviation that we calculated a score for everyone even if under 65 years of age; Australian National University AD Risk Index (ANU-ADRI[7, 8]) with previously adapted year intervals [29]; Cognitive Health and Dementia Risk Index (CogD[8]) and CogD for Alzheimer’s disease (CogD-AD[8]).

Other scores were identified but not included because operationalisation was not possible in the dataset (see Table S2 for a list of algorithms and reasons for exclusion). We only included in this analysis scores whose core risk factors could be proxied in this dataset, accounting for evidence that for some scores not all risk factors are required to conserve index performance[29]. For the selected risk score algorithms, all 1001 participants had complete total scores (see Tables S3-S4 for risk factor weighting, precedent-based adjustments, and observed ranges).

### Pathologic burden

We utilised Amyloid Beta PET (AB PET) positive or negative classification as a recognised gold standard method for diagnosis AD-related pathological burden[30]. A group of experts reviewed the AB PET scans from all sites participating in the original study, providing the positive/negative classification[19]. We did not impute any missing values for this measure, so N = 56 participants were excluded for this outcome.

We utilised pTau-217 from the Lilly Single Molecule Assay (SIMOA), using a previously described distribution-based threshold[31]. We did not impute any missing values for this measure, so N = 181 participants were excluded for this outcome. We chose this more accessible assay over the mass spectrometry University of Gothenburg pTau-217 that was also available in this cohort, to improve generalisability of these findings to clinical settings[32].

We also utilised blood biomarkers for AB40, AB42, total pTau, pTau-181, pTau-217, Glial Fibrillary Acidic Protein (GFAP), Neurofilament Light Chain (NfL), and Soluble Triggering Receptor Expressed on Myeloid cells-2 (STREM2) to explore performance of the scores in explaining difference in a blood-based pathology composite proxy. This analysis is exploratory and reported in Supplementary Materials.

### Statistical analysis

We obtained measures of central tendency of each risk score in the sample and explored discrimination between the clinical and AB PET groups in pairwise comparisons. To that end, we generated Receiver Operating Characteristic (ROC) curves and the associated area under the curve (AUC). We also conducted calibration analysis, using decile calibration plots, Brier scores and Hosmer-Lemeshow tests (calibration slope and intercept are reported in Supplemental Materials).

Initial results indicated significant contribution of the functional item to the performance of the BDSI scale. To investigate this, we also conducted specificity and calibration analysis on the BDSI score generated without including the item “needing help managing medications or money”. To compare the ROC curves for BDSI with and without function, we performed a DeLong test for correlated curves.

P-values are reported exact except where <.001. All analyses were conducted using R in RStudio[33, 34].

## Results

We investigated algorithm performance in the previously described N = 1001 participant cohort of adults aged on average 72 years (SD = 6.57), of which N = 558 (56.5%) were female and N = 143 (14.5%) non-Caucasian. The cohort were classified as cognitively normal (CN, N = 417, 19% from URP), having Mild Cognitive Impairment (MCI, N = 312, 24% from URP), or mild AD (AD, N = 272, 33% from URP). The majority had a negative AB PET scan (N = 603, positive N = 342 that is 36.19% of N = 945 with complete data for this measure). The majority were positive for pTau-217 concentration (N = 469, 57.48% of the N = 816 with complete data for this measure).

We assessed specificity of the scores to four clinical outcome comparisons and two pathology proxies (Table 2 and calibration plots in Supplementary Materials). Only the BDSI achieved clinical level of utility in distinguishing AD from CN (AUC = 0.86). BDSI score also had acceptable performance in distinguishing MCI or AD from CN (AUC = 0.75) and AD from MCI (AUC = 0.71). The only other instances of marginally acceptable specificity were in distinguishing AD from CN using ANU-ADRI (AUC = 0.69), COGD (AUC = 0.69), and COGD-AD (AUC = 0.70, see Supplementary Materials for all values). BDSI was the only risk score to achieve near clinical level discriminant utility for AB PET (AUC = 0.74), similarly outperforming other algorithms for pTau-217 (AUC = 0.67). CAIDE and LIBRA demonstrated chance-level performance for both biomarker-based outcomes.

**Table 1.** Risk factors in each algorithm as implemented in Bio-Hermes-001.

| Risk factor | Risk Index |  |  |  |  |  |
| --- | --- | --- | --- | --- | --- | --- |
|  | CAIDE<sup>1</sup><br>(including APOE e4[26]) | LIBRA [5, 6, 28] | BDSI [18] | ANU-ADRI [7, 8, 29] | CogD [8] | CogD-AD [8] |
| Age | included | - | included | included | included | included |
| Sex | included | - | - | included | included | included |
| Years in education | included | - | included | included | included | included |
| Hypertension | included | included | - | - | included | included |
| BMI | included | included | included | included | included | included |
| High total cholesterol | included | included | - | included | included | included |
| Physical activity | NA | NA | - | - | NA | NA |
| APOE e4 carriership | included | - | - | - | - | - |
| Diabetes | - | included | included | included | included | included |
| Renal dysfunction | - | included | - | - | - | - |
| Coronary heart disease | - | included | - | - | - | - |
| Low/moderate alcohol intake | - | NA | - | NA | - | - |
| Smoking | - | included | - | included | included | included |
| Depression | - | included | included | included | included | included |
| Healthy diet | - | NA | - | - | - | - |
| Cognitive activity | - | NA | - | NA | NA | NA |
| History of stroke | - | - | included | - | included | included |
| Difficulty managing money or medications | - | - | included | - | - | - |
| Traumatic Brain Injury | - | - | - | included | included | included |
| Social engagement | - | - | - | NA | - | - |
| Fish intake | - | - | - | NA | NA | NA |
| Pesticide exposure | - | - | - | NA | - | - |
| Loneliness | - | - | - | - | NA | NA |
| Mediterranean diet | - | NA | - | - | - | - |
| Atrial Fibrillation | - | - | - | - | included | - |
| Insomnia | - | - | - | - | included | - |

*Note.* NA = Not available, meaning typically part of the risk algorithm but not available in the dataset. Dash indicates the variables is not part of a risk algorithm.

**Table 2.**
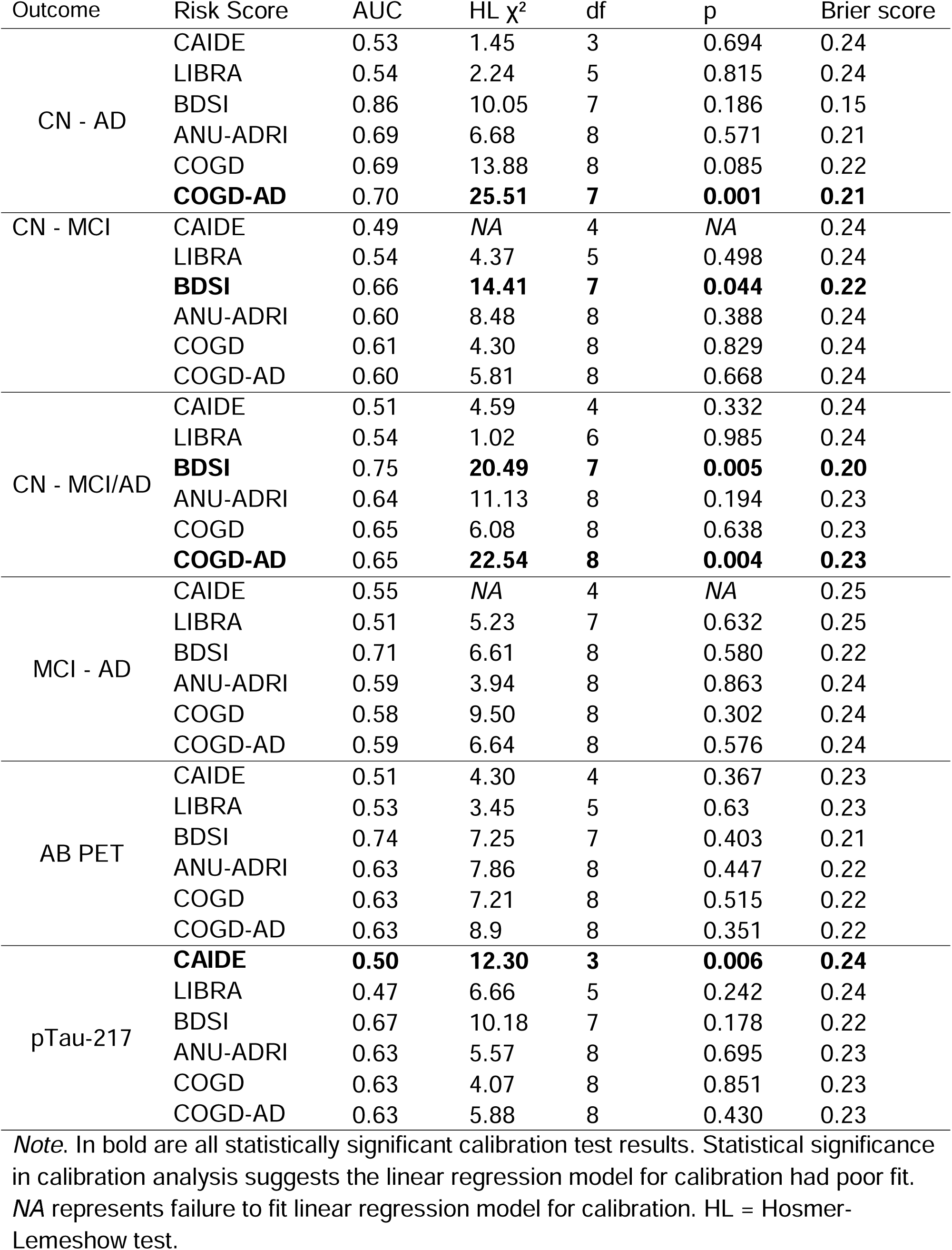
Calibration analysis results for each score and each outcome.

To inform post hoc analysis, we inspected the BDSI risk factor makeup in relation to the other algorithms (see Supplementary Materials for weighting scheme). Functional independence was uniquely incorporated in the BDSI, and where other scores only measured depression based on a Geriatric Depression Scale (GDS) threshold, BDSI also accounted for antidepressants. In this sample, N = 63 participants exceeded the GDS threshold for depression in LIBRA, ANU-ADRI, COGD, and COGD-AD, whereas N = 207 were on antidepressants at the time of study, increasing the total of participants with depression according to BDSI criteria by a factor of approximately 4.29. Although age was accounted for in all assessed scores, BDSI adds a point for every year lived where other weighting schemes are based on age groups.

An ablation study of the contribution of each BDSI item by removing it from the score revealed that removing age produced the lowest correlation to the total score (r = 0.74), followed by function (r = 0.87) and anti-depressants (r = 0.95 compared to removing depression altogether r = 0.96, see also Supplementary Materials). Discrimination analysis supports that the item causing the biggest reduction in AUC was function for clinical phenotypes and biomarker-based comparisons (Table 3, key items also visualised in Figure 1). Calibration analysis (Table 3) showed that removing age led to statistically significantly poor goodness of fit for all tested outcomes even though discrimination remained identical to the total BDSI score. Removing anti-depressants from the BDSI also negatively impacted calibration for distinguishing CN-AD, CN-MCI/AD, and AB PET, whereas removing function only limited calibration for AB PET.

**Figure 1.**
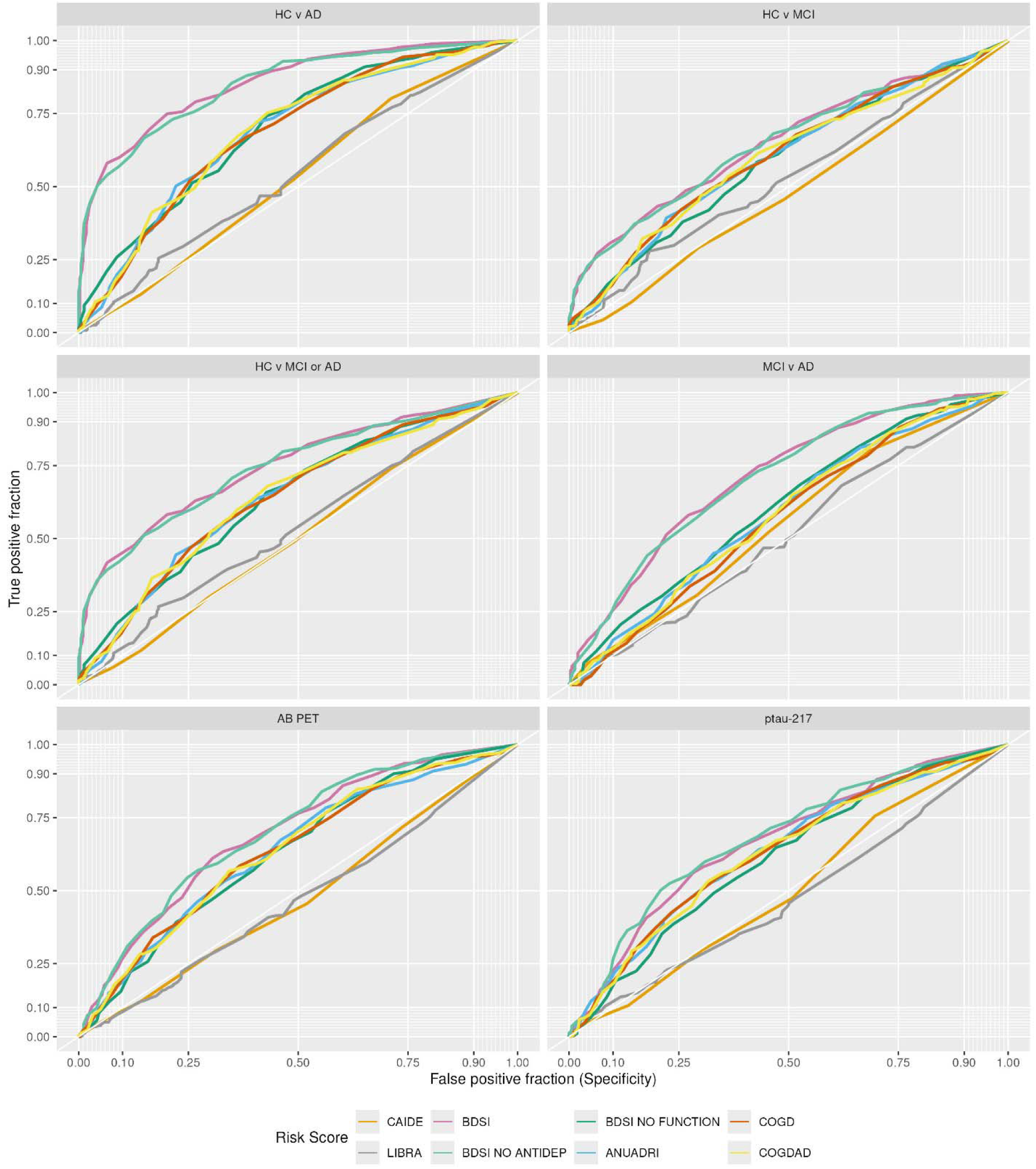
Area Under the Curve (AUC) plots for each algorithm and assessed outcome.

**Table 3.**
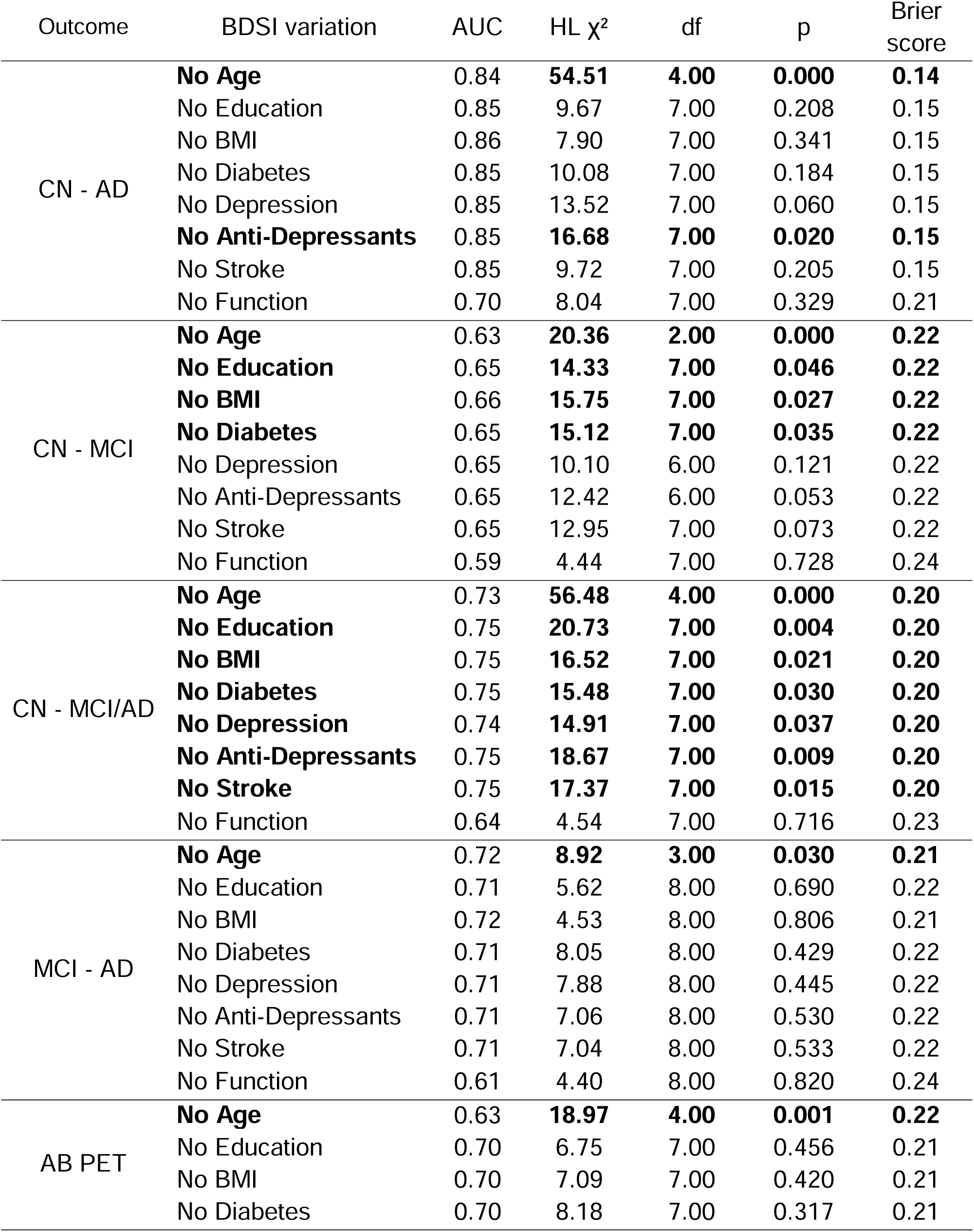
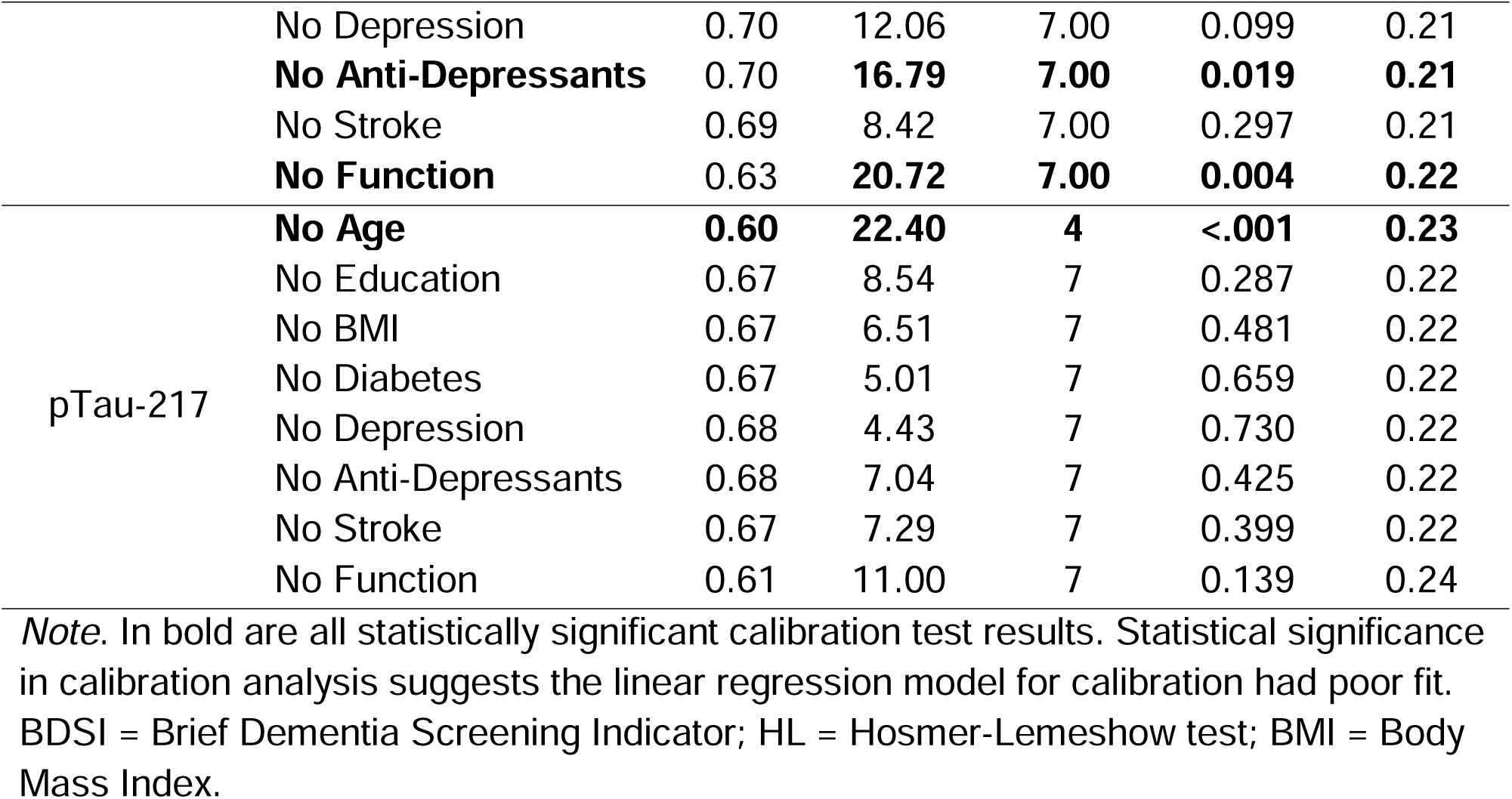
BDSI ablation study discrimination and calibration results.

DeLong test of correlated ROCs evidenced statistically significant differences between the ability of the BDSI with and without function to discriminate all tested clinical and biomarker-based outcomes (Table 4).

**Table 4.** DeLong test of outcome discrimination by BDSI with and without function.

| Outcome | BDSI | BDSI-NF | z | 95% CI | p |
| --- | --- | --- | --- | --- | --- |
| CN - AD | 0.86 | 0.7 | 12.5 | [0.13, 0.18] | <.001 |
| CN - MCI | 0.66 | 0.6 | 7.53 | [0.05, 0.08] | <.001 |
| CN - MCI/AD | 0.75 | 0.64 | 13.1 | [0.09, 0.12] | <.001 |
| MCI - AD | 0.71 | 0.61 | 6.9 | [0.07, 0.13] | <.001 |
| AB PET | 0.7 | 0.63 | 5.87 | [0.04, 0.08] | <.001 |
| pTau-217 | 0.67 | 0.61 | 5.51 | [0.04, 0.07] | <.001 |

*Note.* BDSI-NF = BDSI no function.

## Discussion

In this contemporary, biomarker-characterised cohort, established dementia risk algorithms generally showed limited ability to discriminate current cognitive status as well as amyloid and pTau-217 positivity. Despite acceptable goodness of fit across algorithms, only the Brief Dementia Screening Indicator (BDSI) achieved clinically meaningful discrimination across clinical and biomarker-defined outcomes. An ablation study on the BDSI items evidenced attenuated performance when the functional item was removed. These findings suggest that functional assessment contributes information beyond traditional epidemiological risk factors when distinguishing current clinical and biomarker-defined Alzheimer’s disease.

Importantly, the contribution of the functional item was not restricted to clinical classifications which had accounted for overall functional status. Removal of the item also reduced BDSI discrimination of both amyloid AB PET and pTau-217 positivity, suggesting that the observed effect is unlikely to be explained solely by overlap with diagnostic criteria based on clinical presentation. One interpretation is that functional assessment does not represent another risk determinant. Instead, it represents the integrated downstream consequences of multiple biological, cognitive, behavioural and environmental factors that determine an individual’s ability to independently navigate daily life. This interpretation aligns with other higher-order constructs, like physical frailty[35] or cognitive resilience[36, 37], which are increasingly understood to emerge from domain interactions rather than individual factors.

Although rarely incorporated into AD risk algorithms[38], extended activities of daily living (eADLs), such as managing household tasks beyond ambulating[39, 40], place combined demands on memory, executive function, attention, and physical capacity[41, 42]. As a result, subtle difficulties in independently conducting these activities may emerge before deficits are detectable on standard cognitive tests and may be less dependent on educational attainment, culturally specific knowledge, or other sources of population heterogeneity[43, 44]. In this way, measures of day-to-day cognitive capacity for complex tasks like managing finances with technological aids may capture early cognitive vulnerability that may support dementia screening[45–48], irrespective of determinants of risk that are more distal to lived experience.

Another explanation for the improved discrimination using functional assessment is that the value of risk scores is derived in part from the societal contexts in which they were developed. Access to healthcare, transport, and neighbourhood infrastructure are some of the contextual influences on the extent to which a lifestyle-related risk determinant is truly modifiable within a specific population[49, 50]. Where such structural constraints are common, the clinical utility of lifestyle-related epidemiological risk factors such as BMI may differ compared to populations in which they were developed and previously validated.

Caveating these results are the use of a cross-sectional cohort dataset, without longitudinal follow-up evidence available to support further conclusions regarding utility of the algorithms for risk stratification. Some variables were also adjusted according to precedent (Supplementary Materials provide details in reference to published precedent), which is an inevitable constraint to implementing risk scores in existing real-world datasets. Finally, the BDSI item overlap with the functional activities component of clinical classification presents an important caveat. Although the operational abstraction suffices to facilitate conclusions, its full impact could only ever be ruled out using different instruments.

As such, future directions to consolidate these findings should further validate these analyses using a different source for the BDSI functional item and the clinical classification functional assessment. Similarly, future research should investigate the potential utility of antidepressant use alongside psychometric instrument definitions within dementia risk algorithms, as in the present cohort substantially more participants were identified through antidepressant use than the symptom-based operationalisation. Finally, validation in other datasets, namely longitudinal cohorts, will further consolidate the differences between screening and prognostic algorithms in concurrently stratifying risk.

## Conclusion

In this diverse cohort, most established dementia risk algorithms showed limited ability to discriminate AD-related biomarker positivity and current cognitive status, except for the one algorithm developed to inform referral decisions. Incorporating a functional measure substantially improved discrimination, supporting the diagnostic utility of cognitive vulnerability markers proximal to lived experience. These findings suggest that functional assessment may complement epidemiological risk algorithms in scalable screening and referral pathways as well as cross-sectional investigations.

## Data Availability

All Bio-Hermes-001 cohort data are available on Alzheimers Disease Data Initiative. Access is now open upon request, although that was not the case when this study was initiated.

## Statements

### Ethics

Ethics for access and use of the dataset was provided by the University of Glasgow Medical Veterinary and Life Sciences College Ethics Committee (number 200230198).

### Data availability

The Bio-Hermes-001 dataset (Clinical Trials ID: NCT04733989) is currently hosted on Alzheimer’s Disease Data Initiative (ADDI, report doi: 10.1002/alz.70278)), where it can be accessed upon request (https://discover.alzheimersdata.org/catalogue/datasets/8395cc96-f249-40db-beaa-4668feb3cc5e).

### Code Availability

The authors will be making the risk calculation code for Bio-Hermes-001 openly available on Open Science Framework. The remaining analytical code is not made readily available, but any related queries can be directed to the corresponding author.

### Generative AI statement

In compliance with Wiley guidance for generative AI use, ChatGPT (OpenAI, v 5.5) was used during manuscript preparation to assist with manuscript structure and to improve clarity and readability. During formal analysis, it was also used to streamline looped pipelines. All scientific content was developed by the authors who take full responsibility for the final manuscript.

### Author contributions

KM: conceptualisation; data curation; formal analysis; investigation; methodology; project administration; resources; software; visualisation; writing – original draft; writing – review & editing.

AJD: methodology; software; writing – original draft; writing – review & editing.

LT: methodology; writing – original draft; writing – review & editing.

CD: methodology; writing – review & editing.

JB: methodology; writing – review & editing.

LH: resources; writing – review & editing.

SK: methodology; writing – original draft; writing – review & editing.

TQ: methodology; resources; supervision; writing – original draft; writing – review & editing.

