## supplementary materials for "Functional assessment improves discrimination of clinical and biomarker-defined Alzheimer’s disease"

Functional assessment improves discrimination of Alzheimer’s disease risk over legacy scores: a cross-sectional secondary analysis

**Methods**

**Cohort**

| **Table S1.** Criteria for group classification. | | | |
| --- | --- | --- | --- |
| **Modality of criterion** | **CN (N = 417)** | **MCI (N = 312)** | **Mild AD (N = 272)** |
| Cognition | MMSE score [26, 30]; AND | MMSE score [24, 30]; AND | MMSE [20, 24], or minimum 17 based on clinical judgment |
|  | RAVLT delayed recall score within normal range based on age- and race-adjusted means | RAVLT delayed recall score ≥1SD below age- and race-adjusted means | RAVLT delayed recall score ≥1SD below age- and race-adjusted means |
| Function | No evidence of functional decline based on FAQ score (~0-8) or study partner report, in investigator’s judgment | Minimal to mild functional impairment with preservation of independence in functional abilities based on FAQ score or study partner reports, in investigator’s judgment | Evidence of functional decline and dependence in functional abilities based on FAQ score or study partner reports, in investigator’s judgment |
| Alternative | - | MCI diagnosis based on NIA-AA criteria and verified through medical records | Probable AD diagnosis based on NIA-AA criteria and verified through medical records |

*Note.* Participants with memory scores on RAVLT that exceeded 1.5 SD above age- and race-adjusted mean were not included. CN = Cognitively Normal; MCI = Mild Cognitive Impairment; AD = Alzheimer’s Dementia; MMSE = Mini Mental State Exam^1^; RAVLT = Ray Auditory Verbal Learning Test^2^; FAQ = Functional Activities Questionnaire^3^. NIA-AA = National Institute on Aging – Alzheimer’s Association^4^. Information adapted from^5^.

**Risk scores**

We identified various algorithms to quantifying dementia risk. Some could not be included, as outlined with justification or exclusion in Table S1.

| **Table S2.** Algorithms not included in study. | |
| --- | --- |
| **Algorithm** | **Reason for not including in present study** |
| Ren et al. machine learning model^6^ | Algorithm designed for earlier adulthood, which is outside the age range of the Bio-Hermes-001 cohort. |
| UK Biobank Dementia Risk Prediction (UKDR)^7^ | Algorithm includes variables not available in the Bio-Hermes-001 dataset. |
| Basic Dementia Risk Model (BDRM)^8, 9^ | Algorithm includes variables not available in the Bio-Hermes-001 dataset. |
| Dementia Risk Score^10^ | Variable-level scoring and weighting information not publicly available for the original and UK Bio-Bank adaptation^11^. |
| Short form CogD risk^12^ | Algorithm includes variables not available in the Bio-Hermes-001 dataset. |
| Clinical Dementia Rating (CDR^13^) | Method of administration not compatible with computation in secondary analysis. Variable-level scoring and weighting information not publicly available for use without license for the remotely administered version^18^. |
| Rapid Assessment of Dementia Risk (RADaR)^14^ | Method of administration not compatible with computation in secondary analysis. |

For the scores we did include, we report below detailed adjustments we undertook to implement to the Bio-Hermes-001 dataset, with the scoring and weighting for each risk factor in each algorithm reported in Table S2.

We calculated Cardiovascular Risk Factors, Aging, and the Incidence of Dementia (CAIDE^15^) as previously adapted to include APOE-e4 allele carriership*.* The healthy total cholesterol range originally included 6.5mmol/L^15^ but we adjusted the threshold to be 6.5mmol/L for consistency with the other risk algorithms developed in more recent cohorts. A physical activity measure was missing, but we included APOE carriership (binary yes/no) and used the scoring framework including this variable and its interrelationship with other risk factors^16^. In this dataset, the theoretically possibly maximum CAIDE score could be 17.

We also calculated Lifestyle for BRAin Health (LIBRA^17, 18^) using the Geriatric Depression Scale for depression^19^. We operationalised depression as a score equal to or exceeding 5^20^ on the 15-item Geriatric Depression Scale (GDS), which differs from the 5-item short-form in available scoring frameworks with a threshold of 2 points^21^. Smoking is operationalised in Bio-Hermes-001 as something that had ever occurred before or currently ongoing, which was used as a proxy for the framework’s ongoing/not ongoing approach to the variable. For the framework’s diabetes variable we included both type 1 and type 2, but not any other glucose metabolism disorders. For renal dysfunction, we utilised the broader diagnosis of “renal disorders excluding nephropathies” which included but was not limited to chronic kidney disease. Coronary heart disease variable does not include all coronary artery disorders like coronary artery stenosis. We did not include alcohol because the framework criterion is low/moderate intake per week/day but units or frequency of consumption is not available in this dataset, which only contains information about alcohol consumption being current/past/never. We determined this was not a suitable proxy for alcohol because being a current drinker is insufficient to indicate quantity and would likely introduce error that is harder to quantify than missingness. In this dataset, the theoretically possibly maximum LIBRA score could be 11.6.

We implemented the Brief Dementia Screening Indicator (BDSI^22^) with the deviation that we calculated a score for everyone even if under 65 years of age, although the original publication suggests calculating a score for those over 80. The Bio-Hermes-001 dataset contains embolic stroke, transient ischaemic attack, and thrombotic stroke and basal ganglia stroke which are all categorised as “cerebrovascular accidents” and “central nervous system haemorrhages and cerebrovascular events”, the latter of which includes other conditions too. Instead of using a categorisation from the dataset, we marked every record with the word “stroke” in medical history as having had stroke. For the BDSI item “needing help managing medications or money”, we assigned a positive mark where participants self-reported requiring assistance or being dependent in either one of the following two Functional Activities Questionnaire (FAQ^3^) items: “writing checks, paying bills, balancing check book” and “remembering appointments, family occasions, holidays, medications”. For depression, BDSI framework assigns positive where a patient is taking anti-depressants or self-report “everything was an effort” for 3 or more days in the last week. We determined there was no question in the GDS available in Bio-Hermes-001 that could provide a proxy, so we adjusted the criterion for positive to be taking anti-depressants or GDS score of 5 or more (which is the same threshold we used for CAIDE). In this dataset, the theoretically possibly maximum BDSI score could be 62.

For the BDSI-without function score used in ad-hoc analysis, we retained the entire framework except for the item about “needing help managing medications or money”.

We also calculated Australian National University AD Risk Index (ANU-ADRI^23, 24^) with previously adapted year intervals ^25^. For traumatic brain injury, we used a record of “traumatic brain injury” over the more general category of “cerebral injuries” that can include subdural hematoma, to be in alignment with previous studies^23^. We also followed Anstey et al.^23^ precedent for BMI variable boundaries and to include this algorithm since we had more than 8 risk factors demonstrated to be minimally required for valid performance of the framework. In this dataset, the theoretically possibly maximum ANU-ADRI score could be 68.

Finally, we computed COG-D and COGD-AD: Cognitive Health and Dementia Risk Index (CogD^24^) and CogD for Alzheimer’s disease (CogD-AD^24^). We determined the dataset did not have a suitable proxy for loneliness even through GDS items. For atrial fibrillation, we used the categorisation at higher level that includes paroxysmal atrial fibrillation which are distinct at the lowest level of standardisation. We did the same for insomnia, because higher levels of concomitant condition standardised reporting do not include difficulty sleeping. In this dataset, the theoretically possibly maximum COGD score could be 48 and COGD-AD could be 45.

| **Table S3.** Risk factors, levels, and weights as adapted for Bio-Hermes-001. | | | | | | | | |
| --- | --- | --- | --- | --- | --- | --- | --- | --- |
| **Risk/protective factor** | **Definition in BH** | **Levels and thresholds** | **CAIDE including APOE** | **LIBRA** | **BDSI** | **ANU-ADRI** | **CogD** | **CogD-AD** |
| Age | AGE in DM | <47  [47, 53]  >53 | 0  3  5 | - | - | - | - | - |
|  |  | >= 80  [65, 79] | - | - | no BDSI >80  1 point per year over 65 | - | - | - |
|  | AGE and SEX is female | <65  [65, 70)  [70, 75)  [75, 80)  [80, 85)  [85, 90}  >=90 | - | - | - | 0  1  12  18  26  33  38 | 0  4  7  11  15  19  23 | 0  5  7  13  16  19  23 |
|  | AGE and SEX is male | <65  [65, 70)  [70, 75)  [75, 80)  [80, 85)  [85, 90)  >=90 | - | - | - | 0  5  14  21  29  35  41 | 0  6  8  13  17  20  22 | 0  5  8  12  17  19  23 |
| Sex | SEX in DM | female  male | 0  1 | - | - | - | - | - |
| Years in education | EDUYRNUM in SC | >= 10  (6, 10)  [0,6] | 0  3  4 | - | - | - | - | - |
|  |  | >=12  >6 and <12  <= 6 | - | - | 0  9  9 | - | 0  2  4 | 0  2  4 |
|  |  | >11  [8, 11]  <8 | - | - | - | 0  3  6 | - | - |
| Hypertension | SYSBP in VS | <= 140 mmHg  > 140 mmHg | 0  2 | - | - | - | - | - |
|  | SYSBP and AGE | <= 140 mmHg  > 140 mmHg for AGE <= 65 | - | - | - | - | 0  1 | 0  1 |
|  | SYSBP and DIABP | SBP<140 and DBP <90  SBP>=140 OR DBP>=90 | - | 0  +1.6 | - | - | - | - |
| obesity / being underweight / weight | BMI = WEIGHT/HEIGHT^2 in kg/m^2 and rounded to 2 | <=30 kg/m^2^  >30 kg/m^2^ | 0  2 | 0  +1.6 | - | - | - | - |
|  |  | >= 18.5 kg/m^2^  < 18.5 kg/m^2^ | - | - | 0  8 | - | - | - |
|  | AGE<=65 and BMI = WEIGHT/HEIGHT^2 in kg/m^2 and rounded to 2 | “Underweight” < 18.5  “Normal” [18.5, 25)  “Overweight” [25, 30)  “Obese” BMI >= 30 | - | - | - | -  0  2  5 | 2  0  1  3 | 3  0  1  2 |
| Hypercholesterolemia (hyperlipidemia in code) | (HDL + LDL + .2*TRIG)/18 and rounded to 2 | < 6.5 mmol/L  >= 6.5 mmol/L | 0  1 | 0  +1.4 | - | - | - | - |
|  | AGE < 60 and (HDL + LDL + .2*TRIG)/18 and rounded to 2 | “not high” < 6.5 mmol/L  “high” >= 6.5 mmol/L | - | - | - | 0  3 | 0  3 | 0  3 |
| Physical activity | *not available in this dataset* | NA | NA | NA | - | - | NA | NA |
| APOE e4 status | APOE in C2N (TRUE if E4/x or E4/E4, otherwise FALSE) | non-carrier  carrier | 0  2 | - | - | - | - | - |
| Diabetes | MHHLT “diabetes mellitus included subtypes” | no diabetes  diabetes | - | 0  +1.3 | - | 0  3 | 0  2 | 0  2 |
|  | Low level term MHLLT “Type 2 diabetes mellitus” | no  yes | - | - | 0  3 | - | - | - |
| Chronic kidney disease/renal dysfunction | MHHLGT “renal disorders excl nephropathies)” | no  yes | - | 0  +1.1 | - | - | - | - |
| Coronary heart disease | MHHLGT “Coronary artery disorders” | no  yes | - | 0  +1.0 | - | - | - | - |
| Low/moderate alcohol intake | *No measure of units of alcohol per week in this dataset* | NA | - | NA | - | NA | - | - |
| Smoking | TOBACCO, with SUENRTPT == “ONGOING” in SU | Not currently  currently | - | 0  +1.5 | - | - | 0  1 | - |
|  |  | Never  Former  Current | - | - | - | 0  1  4 | - | 0  0.2  2 |
| Depression | GDS0131 (total) in QS | GDS<5  GDS >= 5 | - | 0  +2.1 | - | 0  2 | 0  3 | 0  4 |
|  | “depression” contained in CMIND in CM ,  GDS0131 (total) in QS | No  Taking antidepressant medications OR  GDS >= 5 | - | - | 0  6 | - | - | - |
| Healthy Diet | *Not available in this dataset* | NA | - | NA | - | - | - | - |
| Cognitive activity | *Not available in this dataset* | NA | - | NA | - | NA | NA | NA |
| History of stroke | MHLLT in MH | No stroke  stroke | - | - | 0  6 | - | 0  2 | 0  2 |
| Difficulty managing money or medications | FAQ0101 (managing finances), FAQ0109 (remembering included medicines) response 2 (requires assistance) or 3 (dependent) | does not require assistance  requires assistance | - | - | 0  10 | - | - | - |
| Traumatic Brain Injury | MHLLT including “traumatic brain injury” in MH | No TBI  TBI | - | - | - | 0  4 | 0  2 | 0  1 |
| Social Engagement | *Not available in this dataset* | NA | - | - | - | NA | - | - |
| Fish intake | *Not available in this dataset* | NA | - | - | - | NA | NA | NA |
| Pesticide exposure | *Not available in this dataset* | NA | - | - | - | NA | - | - |
| Loneliness | *Not available in this dataset* | NA | - | - | - | - | NA | NA |
| Medi-diet | *Not available in this dataset* | - | - | NA | - | - | - | - |
| Atrial fibrillation | AGE > 65 and MHDECOD “Atrial fibrillation” | no  yes | - | - | - | - | 0  2 | - |
| Insomnia | MHDECOD “Insomnia” | no  yes | - | - | - | - | 0  2 | - |

*Note.* NA stands for Not Applicable where a risk factor was not available in the dataset and could thus not be included in calculation. Dash (-) reflects that a risk factor is not included in each column’s risk framework.

| **Table S4.** Range of risk scores by algorithm. | | | | | |
| --- | --- | --- | --- | --- | --- |
| Risk algorithm | Theoretical range | Observed range | Mean (SD) | Median | Observed standardised range |
| CAIDE | [0, 17] | [6, 13] | 8.73 | 8 | [-1.81, 2.84] |
| LIBRA | [0, 11.6] | [0, 9.5] | 2.37 | 1.6 | [-1.32, 3.95] |
| BDSI | [0, 62] | [0, 44] | 12.5 | 12 | [-1.42, 3.57] |
| ANU-ADRI | [0, 68] | [0, 38] | 14.57 | 14 | [-1.52, 2.44] |
| COGD | [0, 48] | [0, 25] | 10.11 | 10 | [-1.87, 2.75] |
| COGD-AD | [0, 45] | [0, 25] | 9.7 | 8.2 | [-1.79, 2.82] |

*Note.* Theoretical range is grounded in the sample characteristics, including maximum age of including participants. These descriptive statistics are provided before standardisation.

**Statistical Analysis**

In an exploratory manner, we also utilised a previously developed and described^26^ principal component unifying blood biomarkers for AB40, AB42, total pTau, pTau-181, pTau-217, Glial Fibrillary Acidic Protein (GFAP), Neurofilament Light Chain (NfL), and Soluble Triggering Receptor Expressed on Myeloid cells-2 (STREM2). Based on their neuropathologic burden proxied in this manner, participants could be arbitrarily clustered according to different criteria. For this analysis we explored two clustering approaches: two clusters reflecting low or high overall pathology; and three clusters reflecting high pTau-dominant pathology, high AB-dominant pathology, and lower overall pathology (for six clusters reflecting high pTau, high AB,

highest AB, and other three groups with pathology of varying types and severity, see Supplementary Materials). We did not impute missing values for this measure, thus N = 11 participants were excluded from both the 2- and 3-cluster pathology models.

Ad hoc analysis of score discrimination in relation to blood-based neuropathology was based on variance explained as captured in Analysis of Variance (ANOVA) omnibus explanatory models. We then visually inspected distributions for pathology clusters as explained by the algorithm with the highest η^2^ and a lower performing one for reference. We did not assess model performance and statistical significance for these ANOVAs that were intended as exploratory tests to contextualise specificity and calibration analysis results in order to inform discussion of future directions with blood biomarkers.

**Results**

| **Table S5.** Calibration analysis results for each score and each outcome. | | | |
| --- | --- | --- | --- |
| Outcome | Risk Score | calibration intercept | calibration slope |
| CN - AD | CAIDE | -2.05 | 4.10 |
|  | LIBRA | -2.07 | 4.15 |
|  | BDSI | -2.68 | 5.33 |
|  | ANU-ADRI | -2.20 | 4.39 |
|  | COGD | -2.18 | 4.34 |
|  | **COGD-AD** | **-2.19** | **2.37** |
| CN - MCI | CAIDE | -2.04 | 4.08 |
|  | LIBRA | -2.04 | 4.09 |
|  | **BDSI** | **-2.15** | **4.30** |
|  | ANU-ADRI | -2.07 | 4.14 |
|  | COGD | -2.09 | 4.18 |
|  | COGD-AD | -2.08 | 4.17 |
| CN - MCI/AD | CAIDE | -2.08 | 4.15 |
|  | LIBRA | -2.08 | 4.15 |
|  | **BDSI** | **-2.31** | **4.52** |
|  | ANU-ADRI | -2.17 | 4.33 |
|  | COGD | -2.18 | 4.35 |
|  | **COGD-AD** | **-2.16** | **4.32** |
| MCI - AD | CAIDE | -2.00 | 4.01 |
|  | LIBRA | -2.00 | 3.99 |
|  | BDSI | -2.22 | 4.42 |
|  | ANU-ADRI | -2.08 | 4.08 |
|  | COGD | -2.02 | 4.03 |
|  | COGD-AD | -2.03 | 4.06 |
| AB PET | CAIDE | -2.15 | 4.38 |
|  | LIBRA | -2.16 | 4.39 |
|  | BDSI | -2.24 | 4.47 |
|  | ANU-ADRI | -2.18 | 4.37 |
|  | COGD | -2.17 | 4.36 |
|  | COGD-AD | -2.18 | 4.37 |
| Ptau-217 | CAIDE | -2.08 | 4.14 |
|  | LIBRA | -2.03 | 4.06 |
|  | BDSI | -2.21 | 4.42 |
|  | ANU-ADRI | -2.15 | 4.29 |
|  | COGD | -2.17 | 4.34 |
|  | COGD-AD | -2.16 | 4.31 |

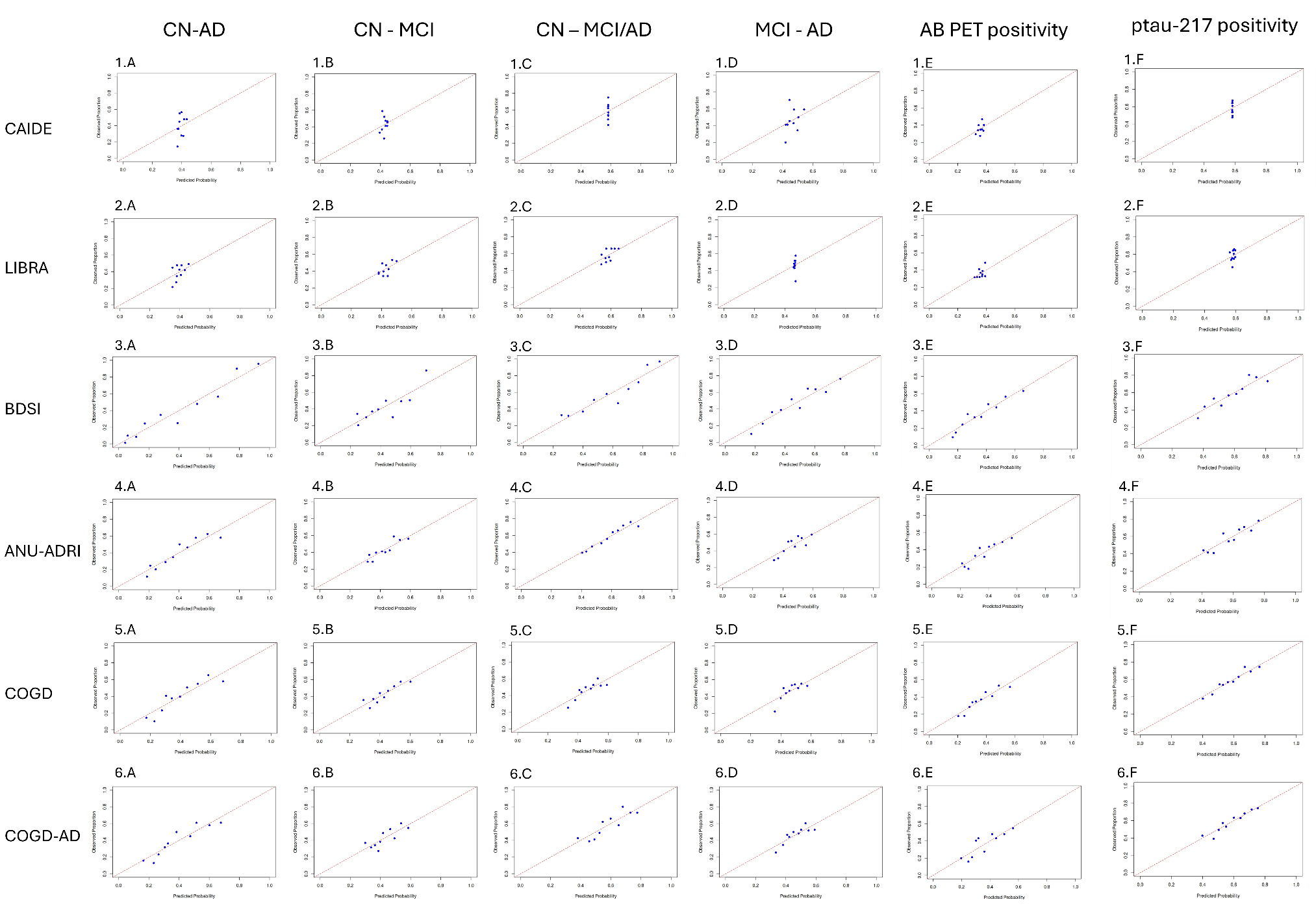
**Figure S1.** Calibration plots for each risk score by clinical outcome assessed.

| **Table S6.** Discrimination and calibration of scores including anti-depressants | | | | | | | | |
| --- | --- | --- | --- | --- | --- | --- | --- | --- |
| Outcome | Risk Score with antidepressants | AUC | HL χ² | df | p | calibration intercept | calibration slope | Brier score |
| CN - AD | LIBRA | 0.55 | 2.59 | 6 | 0.858 | -2.06 | 4.12 | 0.24 |
|  | ANU-ADRI | 0.69 | 9.93 | 8 | 0.270 | -2.20 | 4.38 | 0.22 |
|  | COGD | 0.69 | 10.96 | 8 | 0.204 | -2.17 | 4.33 | 0.22 |
|  | COGD-AD | 0.69 | 8.15 | 8 | 0.419 | -2.20 | 4.37 | 0.21 |
| CN - MCI | LIBRA | 0.52 | 7.09 | 7 | 0.420 | -2.04 | 4.09 | 0.24 |
|  | ANU-ADRI | 0.60 | 3.69 | 8 | 0.884 | -2.07 | 4.14 | 0.24 |
|  | COGD | 0.60 | 9.23 | 8 | 0.323 | -2.09 | 4.18 | 0.24 |
|  | COGD-AD | 0.59 | 9.86 | 8 | 0.275 | -2.08 | 4.17 | 0.24 |
| CN - MCI/AD | LIBRA | 0.53 | 3.46 | 7 | 0.839 | -2.09 | 4.16 | 0.24 |
|  | ANU-ADRI | 0.64 | 11.29 | 8 | 0.186 | -2.16 | 4.32 | 0.23 |
|  | COGD | 0.64 | 8.42 | 8 | 0.394 | -2.16 | 4.32 | 0.23 |
|  | COGD-AD | 0.64 | 4.19 | 8 | 0.839 | -2.14 | 4.29 | 0.23 |
| MCI - AD | LIBRA | 0.54 | 6.82 | 7 | 0.448 | -2.00 | 4.00 | 0.25 |
|  | ANU-ADRI | 0.59 | 6.34 | 8 | 0.609 | -2.04 | 4.07 | 0.24 |
|  | COGD | 0.58 | 11.88 | 8 | 0.157 | -2.02 | 4.05 | 0.24 |
|  | COGD-AD | 0.60 | 10.60 | 8 | 0.225 | -2.04 | 4.08 | 0.24 |
| AB PET | LIBRA | 0.48 | 1.28 | 6 | 0.973 | -2.13 | 4.31 | 0.23 |
|  | ANU-ADRI | 0.64 | 9.54 | 8 | 0.299 | -2.18 | 4.37 | 0.22 |
|  | COGD | 0.64 | 5.91 | 8 | 0.657 | -2.17 | 4.36 | 0.22 |
|  | COGD-AD | 0.64 | 12.01 | 8 | 0.151 | -2.17 | 4.36 | 0.22 |
| Ptau-217 | LIBRA | 0.52 | 8.90 | 6 | 0.179 | -2.01 | 4.01 | 0.25 |
|  | ANU-ADRI | 0.66 | 8.61 | 8 | 0.377 | -2.11 | 4.22 | 0.23 |
|  | COGD | 0.66 | 10.32 | 8 | 0.243 | -2.13 | 4.26 | 0.23 |
|  | COGD-AD | 0.65 | 6.41 | 8 | 0.601 | -2.10 | 4.21 | 0.23 |

*Note.* CAIDE was not included in this analysis as the score does not incorporate depression as a risk factor. HL = Hesmer-Lemeshow test.

**BDSI ablation study**

**Figure S2.** Breakdown of associations between BDSI total and BDSI total with one item removed at a time. Panel A represents an overview of the strength and direction of associations, while panel B demonstrates the distribution of points and correlation value of each pairing.

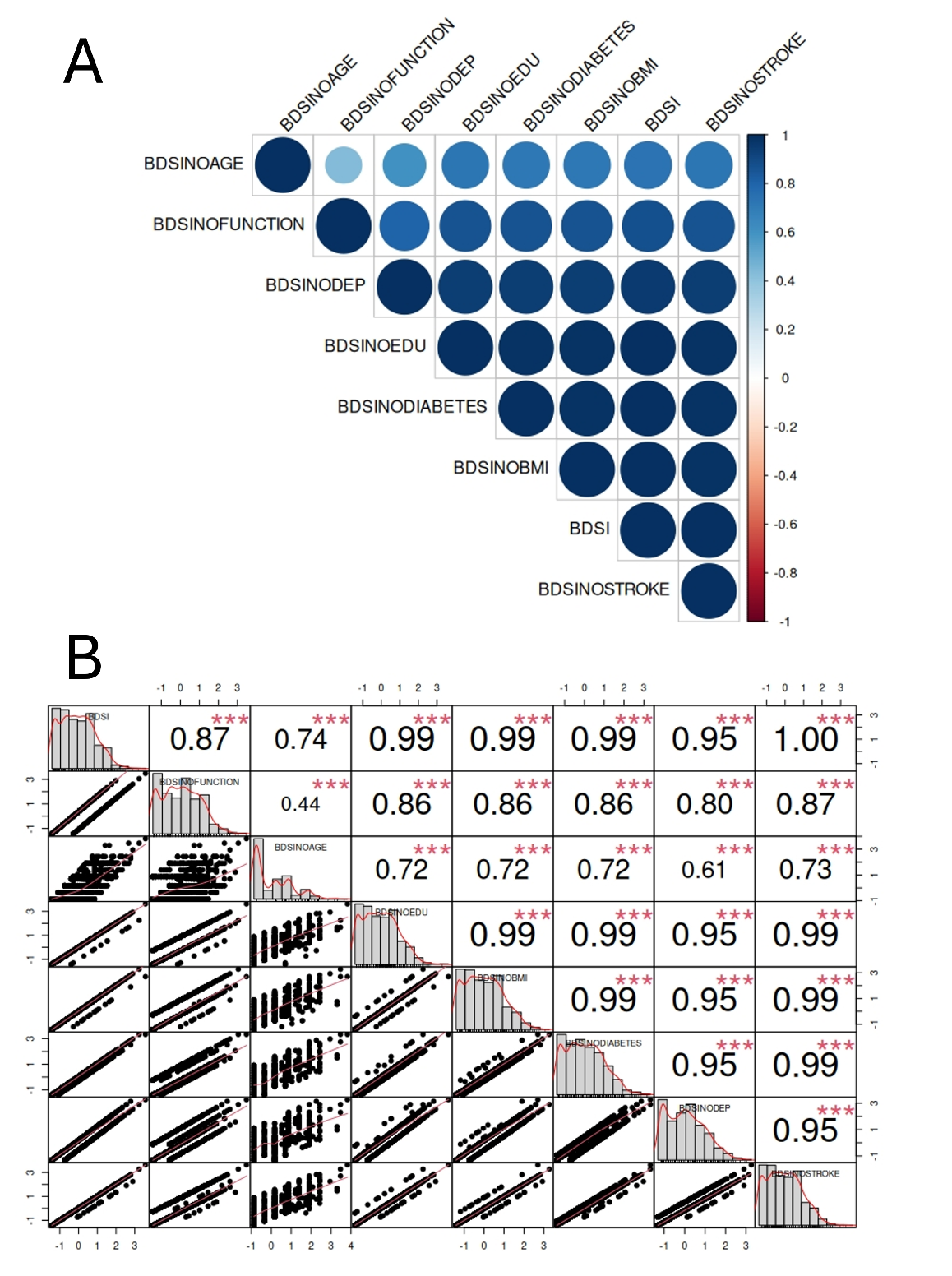

| **Table S7.** BDSI ablation study discrimination and calibration results | | | |
| --- | --- | --- | --- |
| Outcome | BDSI variation | Cal. Int. | Cal. Slope |
| CN - AD | **No Age** | **-2.64** | **5.33** |
|  | No Education | -2.66 | 5.29 |
|  | No BMI | -2.69 | 5.35 |
|  | No Diabetes | -2.67 | 5.31 |
|  | No Depression | -2.62 | 5.23 |
|  | **No Anti-Depressants** | **-2.62** | **5.21** |
|  | No Stroke | -2.63 | 5.24 |
|  | No Function | -2.20 | 4.37 |
| CN - MCI | **No Age** | **-2.13** | **4.29** |
|  | **No Education** | **-2.14** | **4.29** |
|  | **No BMI** | **-2.15** | **4.31** |
|  | **No Diabetes** | **-2.15** | **4.30** |
|  | No Depression | -2.14 | 4.28 |
|  | No Anti-Depressants | -2.14 | 4.27 |
|  | No Stroke | -2.15 | 4.30 |
|  | No Function | -2.07 | 4.13 |
| CN - MCI/AD | **No Age** | **-2.29** | **4.69** |
|  | **No Education** | **-2.22** | **4.49** |
|  | **No BMI** | **-2.24** | **4.54** |
|  | **No Diabetes** | **-2.23** | **4.52** |
|  | **No Depression** | **-2.23** | **4.52** |
|  | **No Anti-Depressants** | **-2.22** | **4.51** |
|  | **No Stroke** | **-2.23** | **4.52** |
|  | No Function | -2.15 | 4.30 |
| MCI - AD | **No Age** | **-2.23** | **4.46** |
|  | No Education | -2.22 | 4.43 |
|  | No BMI | -2.22 | 4.42 |
|  | No Diabetes | -2.21 | 4.41 |
|  | No Depression | -2.20 | 4.40 |
|  | No Anti-Depressants | -2.20 | 4.39 |
|  | No Stroke | -2.20 | 4.40 |
|  | No Function | -2.06 | 4.11 |
| AB PET | **No Age** | **-2.15** | **4.32** |
|  | No Education | -2.25 | 4.49 |
|  | No BMI | -2.24 | 4.47 |
|  | No Diabetes | -2.24 | 4.48 |
|  | No Depression | -2.23 | 4.45 |
|  | **No Anti-Depressants** | **-2.23** | **4.46** |
|  | No Stroke | -2.23 | 4.46 |
|  | **No Function** | **-2.14** | **4.29** |
| Ptau-217 | No Age | **-2.17** | **4.35** |
|  | No Education | -2.19 | 4.38 |
|  | No BMI | -2.19 | 4.40 |
|  | No Diabetes | -2.18 | 4.37 |
|  | No Depression | -2.20 | 4.41 |
|  | No Anti-Depressants | -2.20 | 4.42 |
|  | No Stroke | -2.18 | 4.38 |
|  | No Function | -2.14 | 4.27 |

| **Table S8.** Eta square by risk score | | | |
| --- | --- | --- | --- |
|  | Outcome | | |
|  | Clinical Diagnosis | 2-Cluster pathology | 3-cluster pathology |
| CAIDE | 0.00 | 0.00 | 0.00 |
| LIBRA | 0.00 | 0.00 | 0.02 |
| BDSI | 0.26 | 0.13 | 0.13 |
| BDSI-NF | 0.08 | 0.08 | 0.11 |
| ANU-ADRI | 0.07 | 0.07 | 0.09 |
| COGD | 0.07 | 0.07 | 0.10 |
| COGD-AD | 0.07 | 0.07 | 0.10 |

We also explored the eta square of scores when the cohort is clustered into 6 groups based on pathology (Figure S1). To explore why BDSI was capturing so much more variance than other scores, we visually inspected the distribution of BDSI and COGD scores by each of the 6 pathology clusters (Figure S2), noting that the BDSI scores varied notably between different pathology clusters, for example between the generally high-AB and the highest AB-dominant pathology group. We did not perform ANOVAs on 6-cluster pathology as we did not identify a univariate criterion to distinguish the groups.

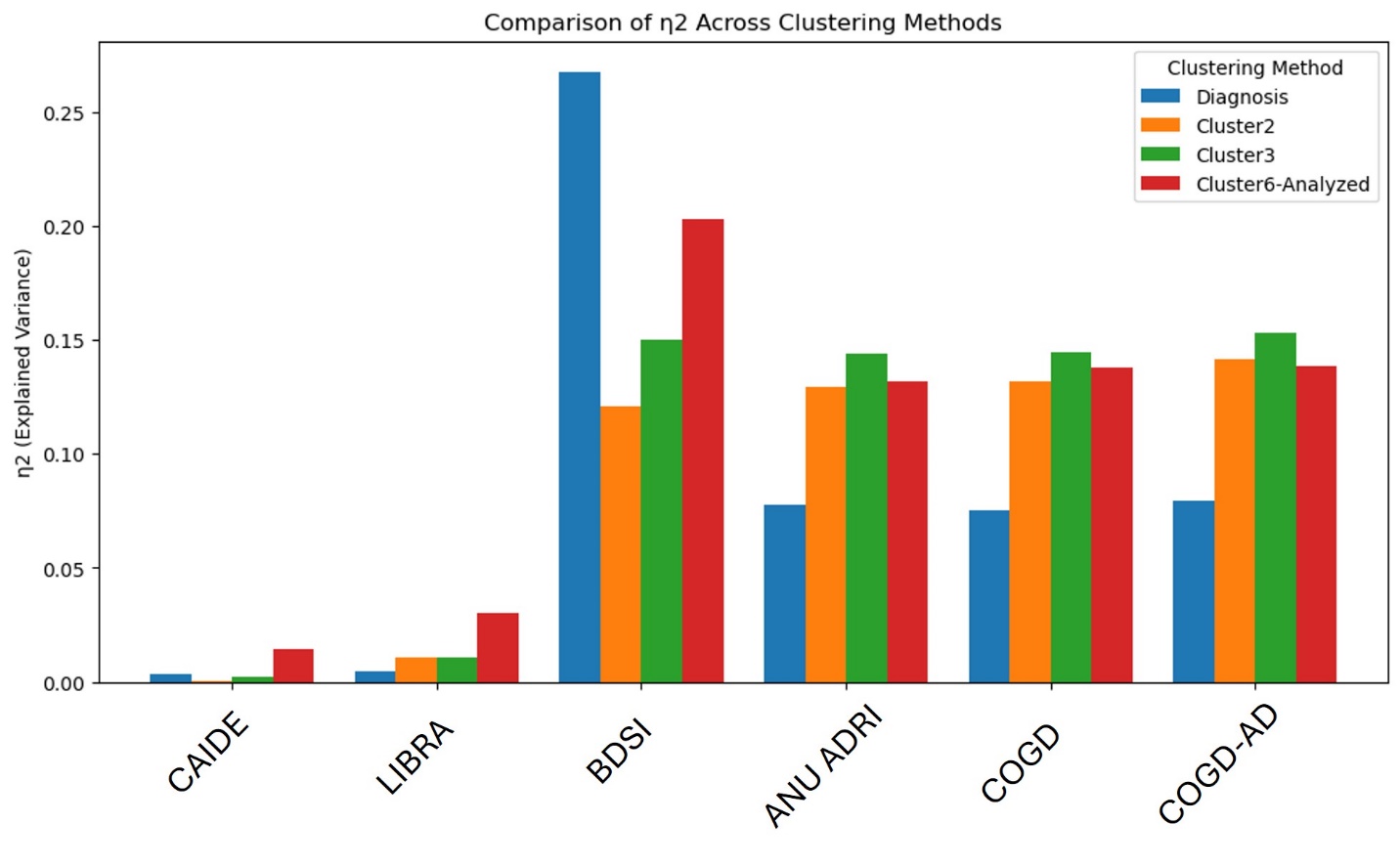

**Figure S3.** Pathology variance explained by different risk scores compared to clinical diagnosis (CN/MCI/AD). Cluster 2 is a binary classification of high/low pathology, whereas Cluster 3 separates the cohort into low/high AB dominant/high pTau dominant.

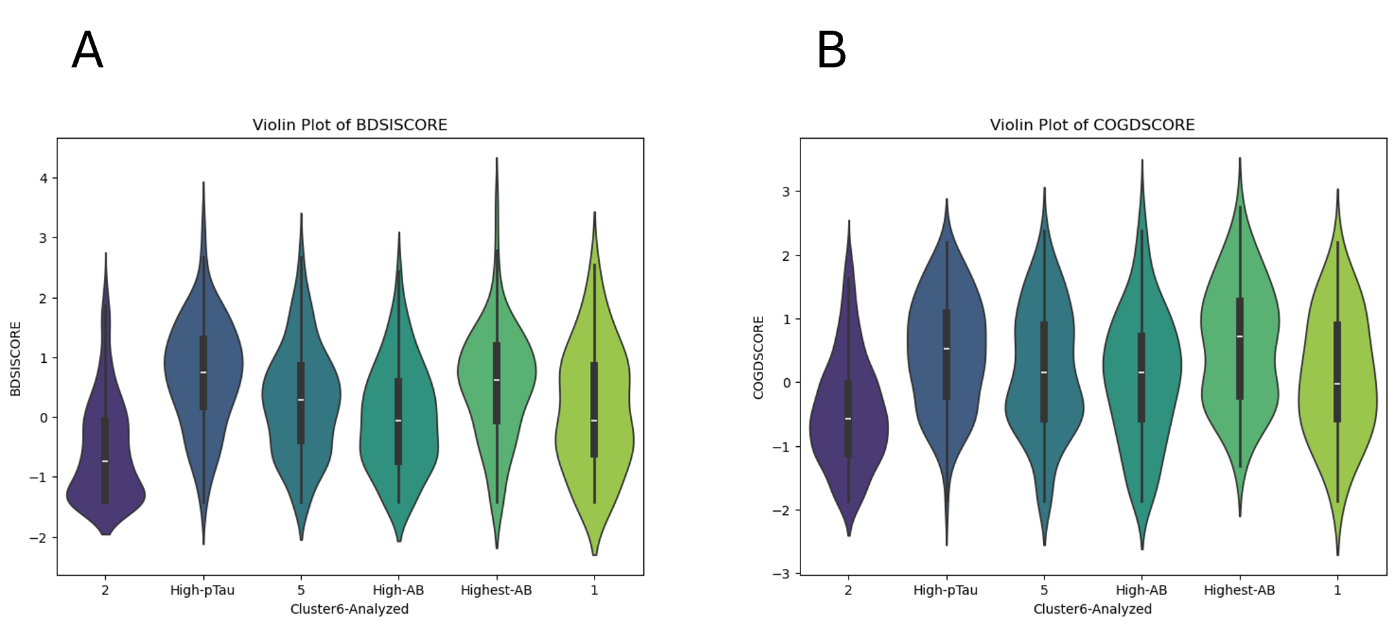

**Figure S4.** Violin plots of score distributions for BDSI (Panel A) and COGD (Panel B) by each of 6 pathology clusters.

**Supplementary Materials references**
